# Long-term effect of empagliflozin and dapagliflozin in patients with heart failure undergoing coronary artery surgery/endovascular intervention: Is there a difference between these two SGLT2i on hospitalization, MACE, and mortality?

**DOI:** 10.1101/2025.02.25.25322846

**Authors:** İlhan Özgöl, Ece Yiğit Gençer, Cennet Yıldız, Dilay Karabulut, Fatma Nihan Turhan Çağlar, Burcu Işıksungur Bıçakhan, Cihan Yücel, Melek Yılmaz, Orçun Ünal, Sadiye Deniz Özsoy, Zerrin Yiğit

**Affiliations:** Department of Cardiovascular Surgery, Prof.Dr. Cemil Taşçıoğlu City Hospital, İstanbul, Türkiye; Department of Internal Medicine, İstanbul Medipol University Faculty of Medicine, İstanbul, Türkiye; Department of Cardiology, Bakırköy Dr. Sadi Konuk Training and Research Hospital, İstanbul, Türkiye; Department of Cardiovascular Surgery, Gaziosmanpaşa Training and Research Hospital, İstanbul, Türkiye; Department of Cardiovascular Surgery, Seyrantepe Hamidiye Etfal Training and Research Hospital, İstanbul, Türkiye; . Department of Cardiovascular Surgery, İstanbul University Cerrahpasa, Faculty of Medicine, İstanbul, Türkiye; . Department of Cardiology, İstanbul University Cerrahpaşa, Faculty of Medicine, İstanbul, Türkiye

**Keywords:** Sodium-Glucose Co-Transporter 2 inhibitors, SGLT2i, empagliflozin, dapagliflozin, coronary artery surgery, heart failure, HFpEF, HFrEF, MACE

## Abstract

Long-term effect of empagliflozin and dapagliflozin in patients with heart failure undergoing coronary artery surgery/endovascular intervention: Is there a difference between these two SGLT2i on hospitalization, MACE, and mortality?

**Objective:** This study aims to compare the long-term effects of these two Sodium-Glucose Co-Transporter 2 inhibitors (SGLT2i) on hospitalization, major adverse cardiovascular events (MACE), and mortality in heart failure patients undergoing coronary artery bypass graft surgery (CABG)/percutaneous coronary intervention (PCI) on empagliflozin and dapagliflozin.

**Methods:** 567 patients with heart failure undergoing CABG/PCI between 2014 and 2022 were studied, and 470 patients on empagliflozin and dapagliflozin were analyzed. The two groups of patients with preserved ejection fraction (HFpEF, n= 293) and patients with low/slightly reduced ejection fraction (HFdEF, n= 177) were analyzed in two subgroups: empagliflozin and dapagliflozin users. In addition to hospitalization, MACE and mortality, age, gender, disease history, and laboratory parameters were compared. Patients with (n=123) and without (n=347) cardiac MACE were also compared.

**Results:** No significant difference was found between the groups in terms of age, HbA1c, creatinine levels, and other cardiovascular risk factors. Similar results were obtained in terms of overall mortality, cardiac mortality, MACE, cardiac MACE, and hospitalization.

- In the HFpEF group comparing dapagliflozin with empagliflozin, overall mortality (17.1% vs. 19.9%, p=0.544), cardiac mortality (10.5% vs. 20%, p=0.341), MACE (29. 6% vs. 26.2%, p=0.522), cardiac MACE (28.3% vs 25.5%, p=0.595) and hospitalization (27% vs 22.7%, p=0.398).
- Similarly, in the HFrEF group, there was no difference in overall mortality (25.9% vs. 13.8%, p= 0.054), cardiac mortality (15.2% vs. 9.2%, p= 0.246), MACE (31.3% vs 20%, p= 0. 105), cardiac MACE (28.6% vs 18.5%, p= 0.134) and hospitalization (28.6% vs. 18.5%, p= 0.134) were similar between the two SGLT2i.

NT-proBNP (1451.38±2769.36 vs. 3052.30±3779.04, p< 0.001) and creatinine (0.98±0.38 vs. 1.12±0.87, p= 0.022) levels were significantly higher in the group with cardiac MACE.

**Conclusion:** Empagliflozin and dapagliflozin did not show a significant difference in their long-term effects on hospitalization, MACE, and mortality in patients with heart failure undergoing coronary artery surgery/endovascular intervention. Larger and multicentre studies are required to confirm these findings.

## Introduction

Diabetes mellitus (DM) is a common disease known to be associated with several complications, in particular atherosclerotic cardiovascular disease (CVD). Effective glycaemic control significantly reduces the risks associated with this disease (1).

While the search for new antidiabetic agents that can effectively control blood glucose levels and significantly reduce the rate of complications continues, the newly launched SGLT2i is encouraging. In addition to being a newly emerging oral antidiabetic, it constitutes a new generation drug group that stands out with its different properties. It is used as a drug to increase glucose excretion by preventing glucose reabsorption in the proximal tubules. It also provides glycaemic control by reducing the amount of glucose in plasma (2).

In addition to glycaemic control, SGLT2 inhibitors have favorable effects on some cardiometabolic markers independent of antidiabetic effects, and their use in cardiovascular (CV) diseases, including heart failure, is beneficial in numerous studies (3,4,5). Even in non-diabetic patients, it has started to be used to prevent adverse effects related to HF, independent of cardiac ejection fraction (EF), whether low or normal (6).

SGLT2i, which effectively reduces CV risks, constitutes a highly preferred drug group today. Compared to conventional oral antidiabetic drugs, empagliflozin and dapagliflozin, which regulate blood glucose without the risk of hypoglycemia, are the two leading agents of this drug group.

Although it has been suggested that there are some differences between SGLT2i, which have been widely prescribed both in terms of antidiabetic and CV system effects, there are few studies on their comparison with each other. In particular, no studies are comparing these two SGLT2i in terms of their long-term effect on hospitalization, major adverse cardiovascular events and mortality in HF patients undergoing CABG surgery/PCI on empagliflozin or dapagliflozin. There appears to be a notable gap in direct comparisons of dapagliflozin and empagliflozin. Consequently, filling this gap to make an optimum contribution is essential by creating an informed decision-making mechanism.

In this study, we aimed to contribute to the existing literature by investigating whether there is a difference between empagliflozin and dapagliflozin differ in terms of the long-term effect of SGLT2i on hospitalization, major adverse cardiovascular events, and mortality in HF patients with CABG surgery/PCI.

## Method

In this retrospective observational study, which covered a follow-up period of 16-23 months, 470 study-eligible patients aged between 26 and 75 years, using empagliflozin and dapagliflozin, were analyzed among 567 HF patients who received SGLT2i and underwent CABG surgery and/or PCI between December 2014 and June 2022.

Patients younger than 18 years of age, older than 75 years of age, diagnosed with type 1DM, uncontrolled hyperglycemia and hypertension, alcohol, drug, and corticosteroid abuse, heart valve replacement, estimated glomerular filtration rate (eGFR) < 60 mL/min/1.73 m 2, and body mass index greater than 30 kg/m2 were excluded.

The study received approval from the Bakırköy Sadi Konuk Training and Research Hospital Ethics Committee (Research Protocol Number: 2025/02, Decision Number: 2025-01-06). All participants gave written informed consent. The research was conducted according to the guidelines of the Declaration of Helsinki.

Blood samples were collected after 10-12 hours of fasting and analyzed by standardized methods on the same instrument in the core laboratory.

All patients underwent echocardiographic examinations at rest according to the guidelines published by the American Society of Echocardiography. Left ventricular ejection fraction (LVEF) was calculated from apical two-chamber and four-chamber volumes obtained at the end of expiration in the left lateral decubitus position using the biplane Simpson technique.

Quantitative evidence of structural and functional cardiac abnormalities consistent with the presence of left ventricular diastolic dysfunction/high left ventricular filling pressures, including elevated natriuretic peptides, symptom +/-finding, and definition of HF by echocardiographic examination, As a result of the LVEF measurement obtained, patients were evaluated and handled in three ways: those with an EF of 50% and above were called HF with preserved EF (HFpEF), those with an EF of 41-49% were called HF with mildly reduced EF (HFmrEF) and those with an EF of 40% or less were called HF with reduced/low EF (HFrEF).

In this study, two groups were formed: patients with preserved EF (HFpEF, Group 1; n= 293) and patients with reduced/slightly reduced EF (HFrEF/HFmrEF, Group 2; n= 177). Each group was subdivided into two subgroups: patients receiving empagliflozin (10 mg) and patients receiving dapagliflozin (10 mg). The number of patients using SGLT2 inhibitors was classified as dapagliflozin (n= 152) and empagliflozin (n=141) in Group 1 and dapagliflozin (n= 112) and empagliflozin (n= 65) and in Group 2.

MACE was defined as a composite of all-cause death, myocardial infarction (MI), stroke, or hospitalization for HF. Cardiac MACE was defined as a composite of death from cardiac causes, MI, and hospitalization for HF. MI was defined as an elevation of at least one cardiac biomarker with a value above the 99th percentile of the upper reference limit, accompanied by ischaemic symptoms or electrocardiographic findings indicating ischemia unrelated to an intervention procedure. A stroke was defined as a neurological deficit lasting more than 24 hours after a sudden cerebrovascular event or acute infarction demonstrated by imaging studies. Hospitalization for HF was defined as hospitalization requiring at least one overnight stay due to significant worsening of HF symptoms and signs, requiring escalation of oral medications or administration of new intravenous HF therapy, including diuretics, inotropes, or vasodilators. Hospitalization for ischaemic heart disease (IHD), typical symptoms and electrocardiographic changes, signs of exercise or pharmacological stress, study evidence for inducible myocardial ischemia, angiographic evidence for new or worsening coronary artery disease (CAD) and intracoronary thrombus, or ischaemic symptoms and signs (electrocardiographic, echocardiographic or biomarker changes) was defined as hospitalization due to the need for coronary revascularisation due to a hospitalization requiring at least one overnight stay due to significant deterioration.

Blood pressure (BP) was measured twice at 10-minute intervals from the left upper arm of patients who were seated and rested for 30-10 minutes and recorded as the mean value. Patients with BP 140/90 mmHg and above were considered hypertensive.

DM, fasting glucose of 126 mg%, and postprandial glycemia equal to or higher than 200 mg% were considered.

The estimated glomerular filtration rate was calculated using the Chronic Kidney Disease Epidemiology Collaboration equation. Chronic kidney disease was defined as eGFR <60 mL/min/1.73 m2, including end-stage renal disease on renal replacement therapy.

The diagnosis of atrial fibrillation (AF) was defined based on the European Society of Cardiology guidelines.

Finally, all patients who underwent CABG surgery and PCI and received SGLT2 inhibitors (n=470) were comparatively analyzed in two groups: patients without cardiac MACE (n=347) and patients with cardiac MACE (n=123).

### Statistics

Continuous variables were expressed as mean and standard deviation, and categorical variables as frequency and percentage. The Kolmogorov-Smirnov test was used to assess whether the data were normally distributed. In comparing two groups, an independent sample t-test was applied if the data were normally distributed, and the Mann-Whitney U test was applied if the data were not. Receiver operating characteristic (ROC)-curve analysis was performed to determine the cut-off value of NT-proBNP for cardiac MACE. Cox regression analysis was performed to determine the independent predictors of cardiac MACE.

### Results

The study included 470 patients with a mean age of 62.89 ± 10.84 years (range: 26-75 years), 59.4% of whom were male. Of the study population, 62.3% had EFpHF, and 37.7% had EFmrHF and HFrEF.

The etiology of HF was ischaemic. The most common comorbidities were hypertension (67.7%), diabetes (64.3%), and atrial fibrillation (24%).

In this study population, 264 (56.2%) patients were on dapagliflozin, and 206 (43.8%) were on empagliflozin. 11.1% of patients were prescribed angiotensin receptor– neprilysin inhibitors (ARNis), 87.2% angiotensin-converting enzyme inhibitors/ angiotensin II receptor blockers (ACEis/ARBs), 72.8% beta-blockers, and 41.3% mineralocorticoid receptor antagonists (MRAs). In addition, 70.4% of patients were taking furosemide and 27.7% were taking torasemide.

In the comparison of the patients in Group 1 with a mean follow-up period of 19.43±1.54 months between the two subgroups formed according to the use of dapagliflozin and empagliflozin, mean age (63.64±10.66 years vs. 61.96±10.50 years, p=0. 175), HbA1c level (6.54±1.36 % vs. 6.68±1.33%, p= 0.314), creatinine level (1.01±0.77 mg/dl vs. 0.92±0.40 mg/dl, p= 0.123), NT - proBNP level (691.80±554.93 pg/ml vs. 574.91±591.03 pg/ml, p=0.155) were similar between the two subgroups. Similarly, no significant difference was observed between the two subgroups regarding HT, DM, AF, smoking, EF, blood urea nitrogen (BUN), uric acid, lipid profiles, and certain cardiac drugs used.

Likewise, when these two subgroups were compared, no significant difference was observed between the two groups in terms of total mortality (17.1% vs. 19.9%, p= 0.544), cardiac mortality (10.5% vs. 14.2%, p= 0.341), MACE (29.6% vs. 26. 2%, p= 0.522), cardiac MACE (28.3% vs. 25.5%, p= 0.595), and hospitalization (27% vs. 22.7%, p= 0.398) in percentage terms. Socio-demographic and clinical characteristics of the patients with ejection fraction over 50% who received dapagliflozin and empagliflozin are shown in **Table 1**.

**Table 1.** Socio-demographic and clinical characteristics of patients with ejection fraction over 50% on dapagliflozin and empagliflozin.

| Variable | DAPAGLIFLOZIN<br>(N=152) | EMPAGLIFLOZIN<br>(N=141) | P-value |
| --- | --- | --- | --- |
| AGE, mean ± SD years | $63.64 \pm 10.66$ | $61.96 \pm 10.50$ | 0.175 |
| Sex, n(%) |  |  | 0.504 |
| Male | 76 (50) | 65 (46.1) |  |
| Female | 76 (50) | 76 (53.9) |  |
| HT, n(%) | 115 (75.7) | 110 (78) | 0.633 |
| DM, n(%) | 106 (69.7) | 85 (60.3) | 0.090 |
| AF, n(%) | 38 (25) | 26 (18.4) | 0.174 |
| Smoking, n(%) | 28 (18.4) | 20 (14.2) | 0.328 |
| EF, mean ± SD | $58.87 \pm 2.86$ | $59.04 \pm 2.69$ | 0.269 |
| ACS, mean ± SD | $119.72 \pm 43.00$ | $123.54 \pm 40.61$ | 0.437 |
| HbA1C, mean ± SD | $6.54 \pm 1.36$ | $6.68 \pm 1.33$ | 0.314 |
| T-C, mean ± SD, mg/dl | $199.50 \pm 49.80$ | $204.69 \pm 54.31$ | 0.406 |
| LDL-C, mean ± SD, mg/dl | $133.54 \pm 44.69$ | $138.46 \pm 49.83$ | 0.381 |
| HDL-C, mean ± SD, mg/dl | $48.20 \pm 15.84$ | $50.96 \pm 14.08$ | 0.038 |
| TG, mean ± SD, mg/dl | $159.30 \pm 113.03$ | $154.64 \pm 89.88$ | 0.850 |
| BUN, mean ± SD, mg/dl | $17.85 \pm 10.80$ | $17.20 \pm 8.14$ | 0.782 |
| Creatinine, mean ± SD, mg/dl | $1.01 \pm 0.77$ | $0.92 \pm 0.40$ | 0.123 |
| Uric acid, mean ± SD, mg/dl | $5.89 \pm 1.85$ | $5.71 \pm 1.65$ | 0.366 |
| NT - proBNP, mean ± SD, pg/ml | $691.80 \pm 554.93$ | $574.91 \pm 591.03$ | 0.155 |
| ARNis, n (%) | 13 (8.6) | 12 (8.5) | 0.990 |
| ACEis or ARBs, n (%) | 139 (91.4) | 133 (94.3) | 0.340 |
| Beta-Blockers, n (%) | 150 (98.7) | 141 (100) | 0.172 |
| MRAs, n (%) | 59 (38.8) | 65 (46.1) | 0.207 |
| Furosemide, n (%) | 108 (71.1) | 101 (71.6) | 0.913 |
| Torsemide, n (%) | 41 (27) | 37 (26.2) | 0.618 |
| OACs, n (%) | 35 (23) | 36 (25.5) | 0.617 |
| CCBs, n (%) | 40 (26.3) | 30 (21.3) | 0.312 |
| Hospitalization, n (%) | 41 (27) | 32 (22.7) | 0.398 |
| Total mortality, n (%) | 26 (17.1) | 28 (19.9) | 0.544 |
| Cardiac mortality, n (%) | 16 (10.5) | 20 (14.2) | 0.341 |
| MACE (total mortality and Hospitalization), n(%) | 45 (29.6) | 37 (26.2) | 0.522 |
| Cardiac MACE, n(%) | 43 (28.3) | 36 (25.5) | 0.595 |
| Follow- up period, mean ± SD months | 19.55±1.62 | 19.30±1.45 | 0.150 |

Abbreviations: HT, Hypertension; DM2, diabetes mellitus 2; AF, Atrial fibrillation; EF, ejection fraction; ACS, acute coronary syndrome; T-C, total cholesterol; LDL-C, low-density lipoprotein cholesterol; HDL-C, high-density lipoprotein cholesterol; TG, triglyceride; BUN, blood urea nitrogen; ARNis, angiotensin receptor–neprilysin inhibitors; ACEis, angiotensin-converting enzyme inhibitors; ARBs, angiotensin II receptor blockers; MRAs, mineralocorticoid receptor antagonists; OACs, oral anticoagulants; CCBs, calcium channel blockers; MACE, major adverse cardiovascular events.

Comparing the patients in group 2 with a mean follow-up of 19.91±1.56 months between the two subgroups formed according to the use of dapagliflozin and empagliflozin, the mean age (63.81±11.67 years vs. 61.57±10.45 years, p=0. 202), HbA1c level (7.41±6.84% vs. 6.67±1.39%, p= 0.895), creatinine level (1.17±0.46mg/dl vs. 1.04±0.39mg/dl, p= 0.051), NT-proBNP (4120.14±4682.70 pg/dl vs. 2785.45±3394pg/dl, p=0.119) level were similar between the two subgroups. Again, no significant differences were observed between the two subgroups concerning HT, DM, AF, smoking, EF, BUN, uric acid, lipid profiles, and certain cardiac medications.

Similarly, when these two subgroups were compared, no significant difference was observed in all-cause mortality (25.9% vs. 13.8%, p= 0.054), cardiac mortality (15.2% vs. 9.2%, p= 0.246), MACE (31. 3% vs. 20%, p= 0.105), cardiac MACE (28.6% vs. 18.5%, p= 0.134), and hospitalization (28.6% vs. 18.5%, p= 0.134). **Table 2** shows the socio-demographic and clinical characteristics of patients with ejection fractions below 50% on dapagliflozin and empagliflozin.

**Table 2.**
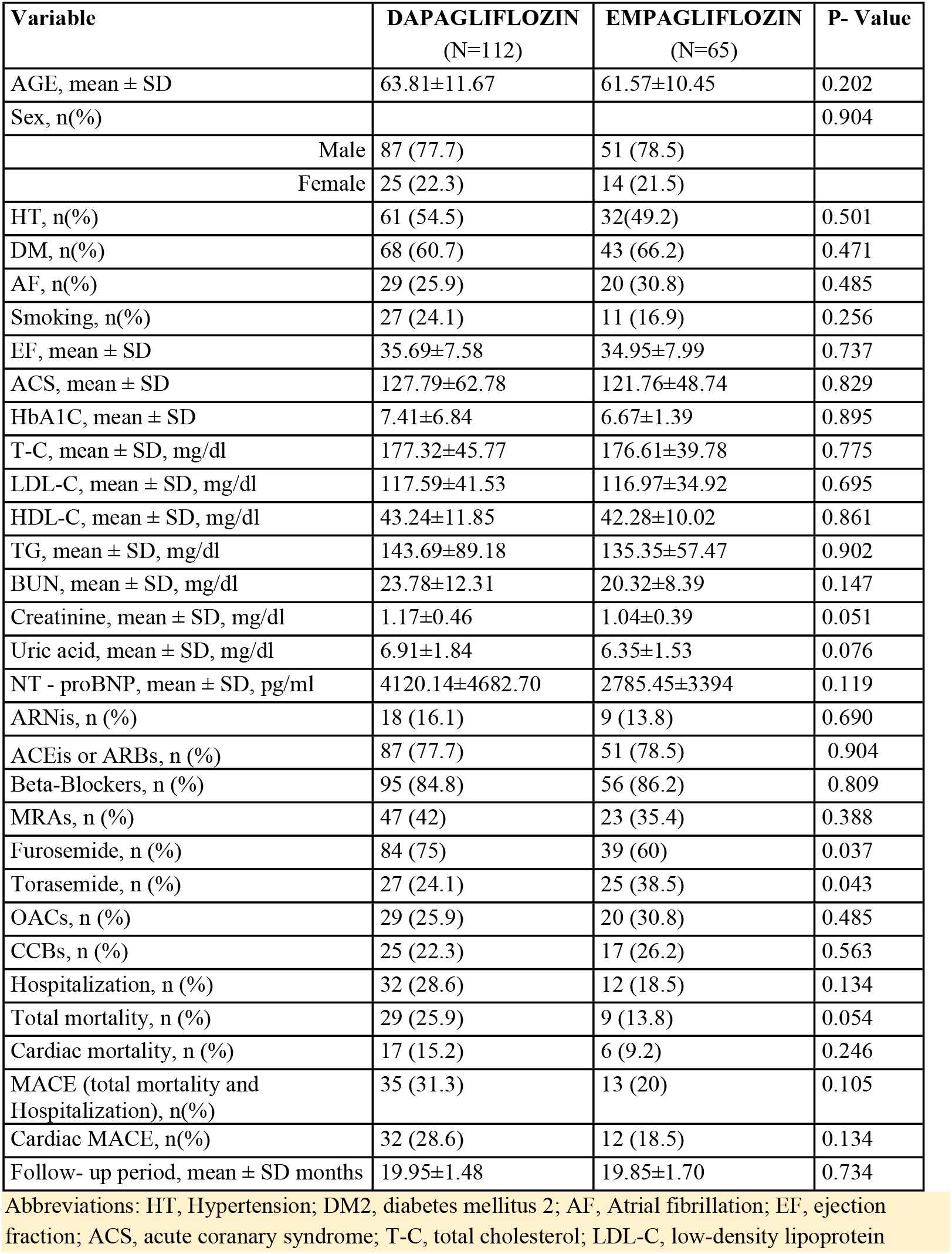
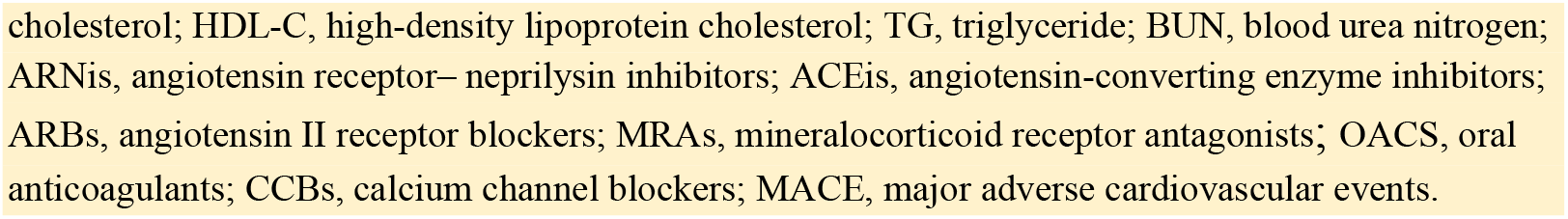
Socio-demographic and clinical characteristics of patients with ejection fraction below 50% on dapagliflozin and empagliflozin.

When all patients were divided into two groups, those who developed cardiac MACE (123) and those who did not develop cardiac MACE (347), and compared in terms of age, gender, DM, AF, smoking, EF, HbA1c, lipid profiles, uric acid, and use of certain cardiac drugs, no significant difference was observed.

The number of patients using SGLT2 inhibitors was similar in both groups, and there was no difference between dapagliflozin and empagliflozin.

A higher proportion of patients with HT was observed in those who did not develop cardiac MACE (70.6% vs. 59.5%, p= 0.022). On the other hand, the number of patients using calcium channel blockers (26.5% vs. 16.3%, p= 0.023) and torasemide (31.1% vs. 17.9%, p= 0.005) was significantly higher in this group.

In the group who developed cardiac MACE, the number of people using beta blockers (98.4% vs. 92.5%, p= 0.018), MRAs (52% vs. 37.5%, p= 0.050), furosemide (83.7% vs. 66%, p< 0.001), and oral anticoagulants (34.1% vs. 22.5%, p= 0.011) was significantly higher than in the other group.

When the group that did not develop cardiac MACE was compared with the group that did develop cardiac MACE in terms of Bun, creatinine, and NT-proBNP levels, significant differences were observed. Mean BUN (18.56±9.35 mg/dl vs. 21.61±0.72, p= 0.002) and creatinine (0.98±0.38 mg/dl vs. 1.12±0.87, p= 0.022) levels were higher in the group of patients who developed cardiac MACE. **Table 3** shows the sociodemographic and clinical characteristics of patients with cardiac MACE and patients without cardiac MACE among all patients on dapagliflozin and empagliflozin.

**Table 3.** Sociodemographic and clinical characteristics of patients with cardiac MACE and patients without cardiac MACE among all patients on dapagliflozin and empagliflozin.

| Variable | Cardiac MACE (–)<br>(N=347) | Cardiac MACE (+)<br>(N=123) | p-value |
| --- | --- | --- | --- |
| AGE, mean ± SD | 63.05±10.96 | 62.59±10.47 | 0.682 |
| Sex, n(%) |  |  | 0.519 |
| Male | 209 (60.2) | 70 (56.9) |  |
| Female | 138 (39.9) | 53 (43.1) |  |
| HT, n(%) | 245 (70.6) | 73 (59.5) | 0.022 |
| DM, n(%) | 227 (65.4) | 75 (61) | 0.377 |
| AF, n(%) | 78 (22.5) | 35 (28.5) | 0.183 |
| Smoking, n(%) | 63 (18.2) | 23 (18.7) | 0.893 |
| EF, mean ± SD | 50.50±11.79 | 49.13±14.35 | 0.876 |
| ACS, mean ± SD | 122.71±48.95 | 124.14±47.73 | 0.879 |
| HbA1C, mean ± SD | 6.66±1.48 | 7.19±6.44 | 0.776 |
| T-C, mean ± SD, mg/dl | 192.95±51.17 | 193.33±49.25 | 0.730 |
| LDL-C, mean ± SD, mg/dl | 130.58±46.27 | 125.76±42.84 | 0.358 |
| HDL-C, mean ± SD, mg/dl | 47.18±13.45 | 47.01±15.98 | 0.543 |
| TG, mean ± SD, mg/dl | 151.81±97.04 | 149.52±89.46 | 0.838 |
| BUN, mean ± SD, mg/dl | 18.56±9.35 | 21.61±0.72 | 0.002 |
| Creatinine, mean ± SD, mg/dl | 0.98±0.38 | 1.12±0.87 | 0.022 |
| Uric acid, mean ± SD, mg/dl | 6.15±1.73 | 6.19±2.02 | 0.736 |
| NT - proBNP, mean ± SD, pg/ml | 1451.38±2769.36 | 3052.30±3779.04 | <0.001 |
| ARNis, n (%) | 35 (10.1) | 17 (13.8) | 0.257 |
| ACEis or ARBs, n (%) | 305 (87.9) | 105 (85.4) | 0.470 |
| Beta-Blockers, n (%) | 321 (92.5) | 121 (98.4) | 0.018 |
| MRAs, n (%) | 130 (37.5) | 64 (52) | 0.050 |
| Furosemide, n (%) | 229 (66) | 103 (83.7) | <0.001 |
| Torsemide, n (%) | 108 (31.1) | 22 (17.9) | 0.005 |
| OACs, n (%) | 78 (22.5) | 42 (34.1) | 0.011 |
| CCBs, n (%) | 92 (26.5) | 20 (16.3) | 0.023 |
| SGLT2i, n(%) |  |  | 0.211 |
| DAPAGLIFLOZIN | 189 (54.5) | 75 (61) |  |
| EMPAGLIFLOZIN | 158 (45.5) | 48 (39) |  |

Abbreviations: MACE, major adverse cardiovascular events; HT, Hypertension; DM2, diabetes mellitus 2; AF, Atrial fibrillation; EF, ejection fraction; ACS, acute coronary syndrome; T-C, total cholesterol; LDL-C, low-density lipoprotein cholesterol; HDL-C, high-density lipoprotein cholesterol; TG, triglyceride; BUN, blood urea nitrogen; ARNis, angiotensin receptor– neprilysin inhibitors; ACEis, angiotensin-converting enzyme inhibitors; ARBs, angiotensin II receptor blockers; MRAs, mineralocorticoid receptor antagonists; OACs, oral anticoagulants; CCBs, calcium channel blockers; SGLT2i, sodium-glucose cotransporter 2 inhibitor.

NT-proBNP (1451.38±2769.36 pg/dl vs 3052.30±3779.04 pg/dl, p< 0.001) levels were significantly higher in the group with cardiac MACE.

Roc Curve analysis of NT-proBNP for cardiac MACE showed that NT-proBNP 2010.50 predicted cardiac MACE with 43.9% sensitivity and 84.2% specificity (AUC: 0.666 P<0.001 CI 95%: 0.580-0.753) (**Fig 1**).

**Fig 1:**
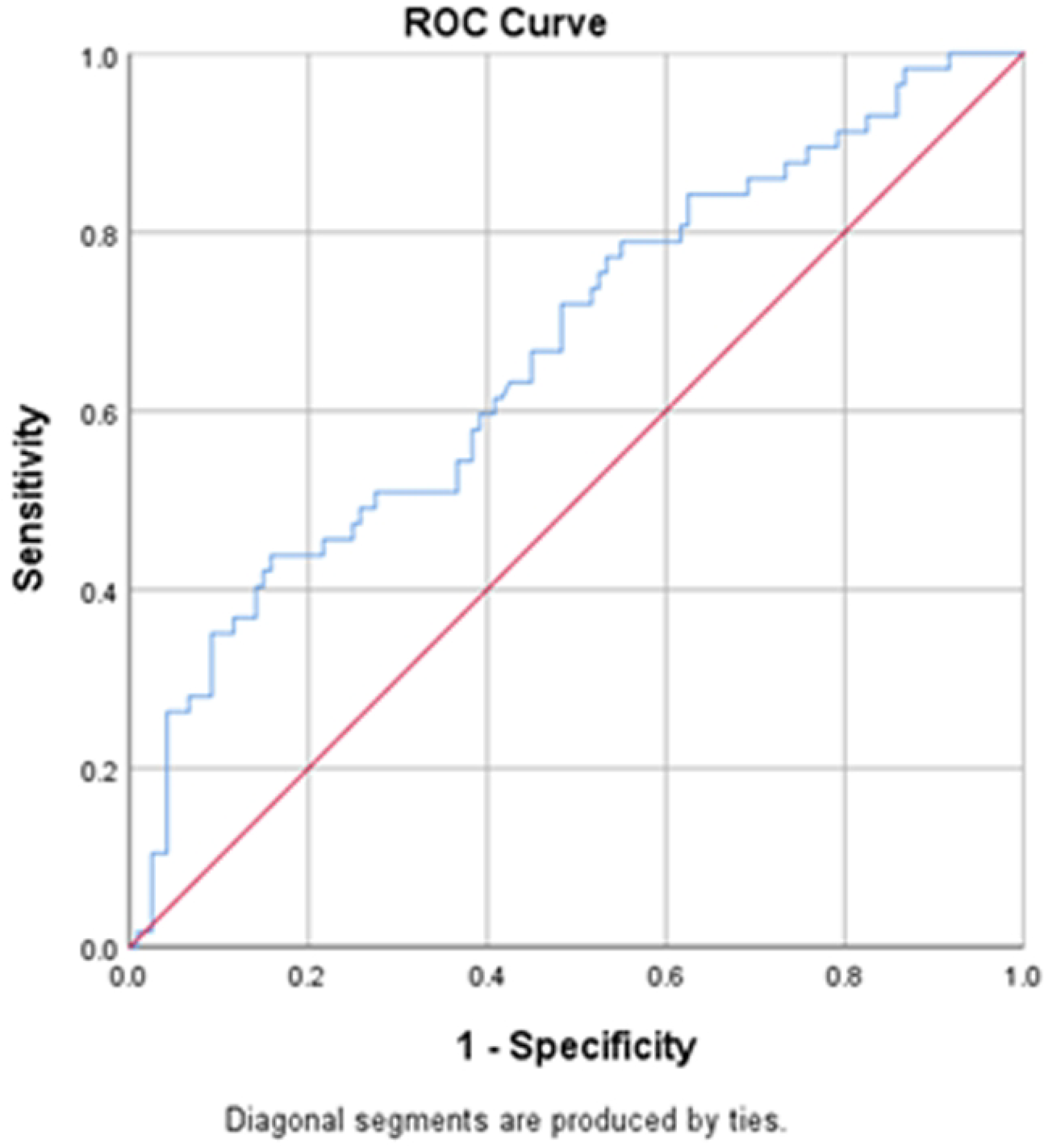
ROC Curve analysis of NT-proBNP for cardiac MACE NT-proBNP 2010.50 Predicts Cardiac MACE with 43.9% sensitivity and 84.2% specifity. AUC: 0.666 P<0.001 CI %95: 0.580-0.753

Cox Regression analysis showed that only NT-proBNP and creatinine predicted cardiac MACE in patients receiving SGLT2i (P: 0.001 for NT-proBNP, HR: 1.023, CI: 1.012-1.123 and for creatinine P: 0.046, HR: 1.529, CI: 1.007-2.322) (**Table 4**).

**Table 4.** COX Regression of NT-proBNP and creatinin for cardiac MACE.

|  | **p** | **HR** | **CI** |
| --- | --- | --- | --- |
| **NT-proBNP** | 0.001 | 1.023 | 1.012-1.123 |
| **Creatinine** | 0.046 | 1.529 | 1.007-2.322 |

Only NT-proBNP and creatinine predict cardiac MACE in patients receiving SGLT2i.

Finally, survival according to NT-proBNP is shown in **Fig 2**.

**Fig 2:**
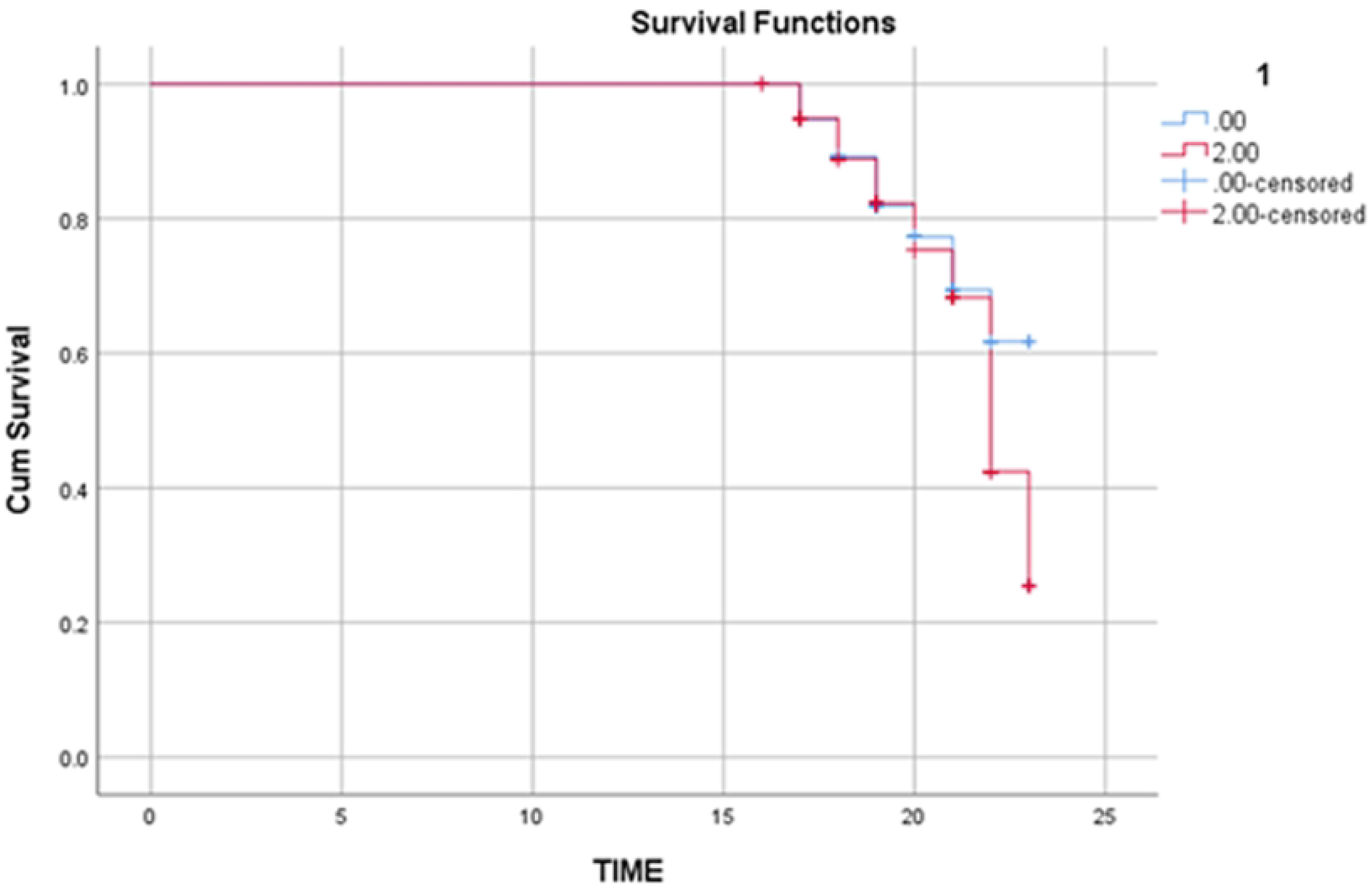
Survival according to NT-proBNP

## Discussion

SGLT2i is known to reduce CV mortality and HF-related hospitalization rates in patients with HF (2,7). Although significant progress has been made in providing optimal treatment for managing cardiovascular risk, the issue of selecting the appropriate SGLT2i has not been adequately clarified.

Aside from the fact that the comparative results of empagliflozin and dapagliflozin in HF patients using SGLT2i are scarce and poorly defined, it is clear that there are no studies comparing empagliflozin and dapagliflozin in HF patients undergoing CABG surgery and PCI and using SGLT2i. Based on this, in the present study, there was no significant difference between empagliflozin and dapagliflozin in terms of total mortality, cardiac mortality, MACE, cardiac MACE, and hospitalization when comparing both subgroups, whether in the preserved EF group or the low/mildly reduced EF group.

In this study comparing SGLT2 inhibitors in the preserved or low/mildly reduced EF group, mean age, HbA1c, creatinine, and NT—proBNP levels were similar between empagliflozin and dapagliflozin, with no significant difference between them. On the other hand, HT, DM, AF, smoking, fasting blood glucose, BUN, uric acid, lipid profiles, and certain cardiac medications were similar between both subgroups.

When considered, it can be said that our present study is in parallel with some studies that showed no significant difference between empagliflozin and dapagliflozin in terms of CV outcomes and which compared SGLT2i in a patient population including not only coronary artery patients who underwent CABG surgery and PCI but also all DM patients with HF in general (8,9).

In a recent single-center retrospective observational study in which 1549 patients who were prescribed empagliflozin/dapagliflozin were examined, Jee-Heon Kim et al. suggested that there was no significant difference between empagliflozin and dapagliflozin in terms of MACE occurrence as a result of their 3-year examination. In addition, it was emphasized that there was no significant difference between the two groups regarding all-cause mortality, CV mortality, MI, hospitalization for HF, or cerebrovascular events (10).

On the other hand, when analyzed, it is also seen that there are opposing studies showing a difference between empagliflozin and dapagliflozin. As in a multicentre retrospective cohort study that sought to answer the question ‘What are the comparative results of empagliflozin and dapagliflozin in reducing all-cause mortality and hospitalizations in patients with HF?’, Katherine L. et al. in their 1-year comparative study of 28075 patients with HF who had never received SGLT2i and who had just started empagliflozin/dapagliflozin treatment, it was emphasized that patients on empagliflozin had a lower probability of all-cause mortality or hospitalization compared with patients on dapagliflozin(11).

In 14 randomized controlled meta-analysis studies involving 75334 patients investigating the effect of SGLT2i on patient outcomes, Chen JY et al. analyzed 40956 patients receiving SGLT2i and suggested that empagliflozin users with diabetes had a significantly lower risk of all-cause mortality compared with dapagliflozin users(12).

In another meta-analysis of 47 randomized controlled trials involving 70574 patients, Jiang Y et al. suggested that once-daily empagliflozin (10 mg/25 mg) may be better than dapagliflozin and other SGLT2i because of its lower risk of all-cause mortality and CV events in T2DM patients (13).

On the other hand, some studies conclude that dapagliflozin is superior to empagliflozin compared to empagliflozin. One of these is a network meta-analysis study that included 11 randomized controlled trials comparing the efficacy of dapagliflozin, empagliflozin, and placebo in heart failure patients (14). This study by Zepeng Shi et al. stated that dapagliflozin was superior to empagliflozin and placebo in reducing mortality due to all causes and cardiovascular causes.

In this study, it was also reported that dapagliflozin was equivalent to empagliflozin in terms of hospitalisation for HF and HF-related mortality/hospitalisation and had a significantly lower effect on HF exacerbation with empagliflozin compared to placebo. However, when compared with empagliflozin, dapagliflozin had a significantly lower effect on HF exacerbation.

From a different perspective, when all patients in our study were divided into two groups, those who developed cardiac MACE and those who did not develop cardiac MACE, and compared in terms of age, gender, DM, AF, smoking, EF, fasting blood glucose, HbA1c, lipid profiles, uric acid, and use of certain cardiac drugs, no significant difference was observed. In the comparison between the groups, patients on SGLT2i were similar in both groups, and there was no difference in the use of dapagliflozin and empagliflozin. However, when these two groups were compared in terms of bun, creatinine, and NT-proBNP levels, it was observed that there were significant differences between them; the mean bun and creatinine levels were higher in the group of patients with cardiac MACE, especially NT-proBNP levels were significantly higher in the group with MACE.

Roc Curve analysis of NT-proBNP for cardiac MACE showed that NT-proBNP predicted cardiac MACE. Furthermore, Cox Regression analysis showed that NT-proBNP and creatinine predicted cardiac MACE in patients receiving SGLT2.

The limitations of this study were that the duration of time from the onset of the first heart failure to treatment was not known; the variability of the follow-up period may affect the clinical results; the fact that not all patients were offered each component of the 4-component treatment regimen recommended for the optimal treatment of heart failure in light of current guidelines; the lack of information on patients’ compliance, lifestyle factors, and dietary habits; the fact that it was a single-center retrospective observational study; the relatively small sample size; and the lack of a control group.

Studies with a larger sample size and the inclusion of a control group should be conducted to increase the statistical power and ensure the reliability and generalizability of the findings.

## Conclusion

Our study showed that there was no difference between empagliflozin and dapagliflozin in the comparative long-term effect of these two SGLT2i on hospitalization, major adverse CV events, and mortality in HF patients on SGLT2i undergoing CABG surgery and PCI. However, more extensive studies are needed to confirm and generalize these findings, and large multicentre observational or randomized controlled trials are needed.

## Data Availability

There is no limitation. But, the data may be kept in a public repository and only available after acceptance. The authors' contact numbers, DOIs, etc. If additional information is requested, I kindly request that I be informed about this issue.

## Notes

### Competing Interest Statement

The authors have declared no competing interest.

### Funding Statement

The author(s) received no specific funding for this work.

### Author Declarations

The study received approval from the Bakırköy Dr. Sadi Konuk Training and Research Hospital Ethics Comitee (Research Protocol nNumber: 2025/02, Decision Number: 25-01-06).

